# Antidepressant prescribing trends for adults with and without autism in the United Kingdom from 1997 to 2023, a population-based cohort study using Clinical Practice Research Datalink Aurum

**DOI:** 10.1101/2025.10.24.25338725

**Authors:** Aws Sadik, Golam M. Khandaker, Antonio F. Pardinas, Michael Lundberg, Brian Lee, Cecilia Magnusson, Dheeraj Rai, Paul Madley-Dowd

## Abstract

**Background:** Antidepressant prescribing and autism diagnoses have both increased in recent years, however antidepressant prescribing patterns for autistic adults are not well understood. Aims: (1) To describe antidepressant trends in autistic and non-autistic adults in the UK from 1997 to 2023. (2) To compare trends between autistic adults and age- and sex-matched non-autistic adults. (3) To investigate how trends vary by intellectual disability (ID) status, age-group and sex.

**Methods:** Using population-representative primary care records from the Clinical Practice Research Datalink Aurum, we defined three autistic groups - autistic adults with ID, autistic adults without ID and all autistic adults - and four non-autistic groups - all non-autistic adults and an age- and sex-matched comparator group for each autistic group. In each calendar year we calculated: annual, lifetime and new antidepressant prescribing; indications for serotonin-selective reuptake inhibitor initiation; average doses prescribed for citalopram, fluoxetine, and sertraline; and the proportion of courses that lasted over 1, 2 and 3 years. We performed primary analyses in 16-64 year olds and repeated them with stratification by sex and age-group.

**Results:** In all, 34,173,295 non-autistic adults and 172,242 autistic adults were included (47,011 with ID and 125,231 without ID). 30% of autistic adults and 14.7% of non-autistic adults were prescribed an antidepressant in 2023. Annual prescribing, average doses and course durations increased for all groups during the study period, and were higher for autistic adults than non-autistic adults. Annual, new and lifetime prescribing were highest in autistic adults without ID, whereas course durations were highest in autistic adults with ID. Recording of depression and anxiety as indications was lower for those with ID. Trends were largely similar across sex and age strata, with higher prescribing among females.

**Conclusions:** Antidepressant prescribing for autistic adults has increased since 1997 and patterns of use vary by intellectual disability status.

## Introduction

Autism is a neurodevelopmental condition characterised by social communication difficulties and restricted and repetitive behaviours [1]. Antidepressants are among the most commonly prescribed medications in the United Kingdom (UK), used by approximately 15% of adults in England [2]. They can be prescribed for conditions that commonly co-occur with autism, such as anxiety and depression, and have also been used off-licence for features of autism that some autistic people find distressing, such as repetitive behaviours [3]. Antidepressant prescribing in the UK has increased over several decades with a concurrent increase in long-term maintenance prescriptions[4, 5], heightening scrutiny on whether benefits and harms are being appropriately reviewed for patients.

NHS England has highlighted potential over-prescribing of psychotropic medications for autistic people [6] and there is some evidence that antidepressant prescribing has been high [7–10] but these studies have involved mostly children and performed limited comparisons to non-autistic groups. There are limited contemporaneous data on antidepressant prescribing patterns for autistic adults, how these have changed over time, and how these compare to non-autistic adults. Furthermore, it is unclear whether there are systematic differences in the reasons antidepressants are used, what doses are prescribed, and for how long they are continued. Each of these is likely to vary by age and sex, given prescribing prevalence differences seen in the general population [5]and sex differences in co-occurring conditions among autistic people [11].

To fill this gap in the literature, we used population-representative UK primary care records towards the following aims: (1) To describe antidepressant prescribing trends in autistic and non-autistic adults from 1997 to 2023. (2) To compare these trends between autistic adults and age- and sex-matched non-autistic adults. (3) To investigate how these trends vary by intellectual disability status, age-group and sex. Specifically, we sought to characterise annual prescribing, new prescribing, lifetime history of prescribing and long-term prescribing of antidepressants. We also sought to characterise the recorded prescribing indications for the most commonly used antidepressant class, serotonin-selective reuptake inhibitors (SSRIs), as well as average doses prescribed for the three most common SSRIs.

## Methods

### Design/Setting/Data sources

The study protocol was pre-registered and updated at https://osf.io/em9gw/ [12] and a RECORD-PE checklist is in the supplement [13].

We performed a descriptive cohort study set between 1^st^ January 1997 and 31^st^ December 2023, using the March 2024 extract of the United Kingdom Clinical Practice Research Datalink (CPRD) Aurum database [14]. CPRD Aurum contains routinely-collected electronic health records from primary care practices that use EMIS Web records and is representative of the English population in age, sex, geographical spread and deprivation [15]. The March 2024 extract included 47,413,279 research “acceptable” patients from 1,784 general practices [14]. “Acceptable” patients are flagged by CPRD as having research-quality data based on the recording and internal consistency of key variables [15].

### Ethics & Consent Statements

The authors assert that all procedures contributing to this work comply with the ethical standards of the relevant national and institutional committees on human experimentation and with the Helsinki Declaration of 1975, as revised in 2013. All procedures involving human subjects/patients were approved by the Independent Scientific Advisory Committee of CPRD (protocol number 23_002605). Individual patient consent for this study was not required as all data sent to CPRD by GP practices is pseudonymised at source, with patients able to opt-out of data sharing for research purposes.

### Participants

The eligible patient list was derived from a CPRD database denominator list that included information on year of birth, registration date, registration end, death date and practice last data collection date. Individuals were eligible for inclusion to analyses if the following criteria were met: completed 1 year of registration at a general practice on 1st January 1997 or later; have the CPRD ‘acceptable’ data quality flag (present for over 93% of registrations [14]); 16 years of age or older (the age at which transition to adult health services begins in the UK). Full date of birth is not held by CPRD to protect patient confidentiality, so date of birth was assumed to be the first day of the year of birth. Individuals were followed-up until the end of their registration at the practice; the last data collection date for their practice, their death, or 31st December 2023, whichever came first.

The cohort was divided by the lifetime presence or absence of autism and intellectual disability (ID) diagnostic codes to create three groups: autistic adults with ID, autistic adults without ID, and non-autistic adults. Autistic adults with and without ID were also combined to form an overall autistic group.

Matched comparator groups were also prepared for each of the three autistic groups. For each autistic adult we identified up to four non-autistic adults that were matched on year of birth, sex, and the year in which they became eligible to enter the study.

### Variables and codelist development

Covariates included sex, ethnicity, region and mental health variables. The mental health variables identified for comparing group characteristics were: attention deficit hyperactivity disorder (ADHD); depressive disorders (depression); anxiety and fear-related disorders (anxiety); self-harm; and records of mental health contact, referral or signposting (psych contact). Mental health variables used to assess potential prescribing indications were: depression; anxiety; behaviours that challenge; and other mental health conditions (post-traumatic and stress-related disorders; somatoform and dissociative disorders; obsessive-compulsive and related disorders; eating disorders and personality disorders). Antidepressants included in the study, and their categories, are listed in supplementary table 1.

The CPRD Code Browser was used by AS (a psychiatric resident doctor) to prepare mental health and ethnicity code lists with cross-checking against relevant published lists [[16]][17][[18]][19][20]. Where relevant, advice from another clinician (DR) was sought. Further details are provided in the supplementary methods.

### Prescription duration and dose cleaning

We compiled information on each drug prescription and dose for the cohort during the study period. The duration of a prescription was imputed if the recorded duration was zero days (i.e. missing) or greater than 185 days (i.e. considered implausibly long). Duration was imputed by dividing the quantity prescribed by the daily dose prescribed, as long as the units for these values matched. If these variables were also missing, the duration was set as missing. The approach to cleaning of dose information varied depending on whether multiple issues of the same substance were prescribed for someone on the same day, with full details provided in the supplementary methods.

### Analyses

#### Primary analyses

We performed primary analyses in 16-64 year-olds inclusive, because prescribing guidelines often differ for over 65s, and preparatory analysis of the cohort indicated that the proportion of over 65s among autistic adults was very low. All analyses were stratified by calendar year to identify temporal trends. In the analyses of trends over time, no matching or standardisation of the non-autistic group was performed, so that results reflected trends in non-autistic 16-64 year olds as a whole including demographic changes. Then, to allow comparisons between autistic adults and non-autistic adults with less risk of bias due to confounding by age and sex, we repeated the analyses with three matched non-autistic comparator groups, as described below.

#### Descriptive analysis

For each comparator group we reported the counts and percentages of people in each category of sex, age group, cohort entry period, region, and ethnicity, and median follow-up time from cohort entry to exit. We also reported the counts and percentages of people in each group with mental health conditions recorded at any time prior to cohort exit. This was also repeated with restriction to patients included in calendar years 1997, 2010 and 2023.

#### Annual prescriptions, lifetime prescriptions and new prescriptions

The annual proportion prescribed antidepressants were calculated as the proportion of individuals in receipt of a prescription in a calendar year, among those with full follow-up for that calendar year. This was performed for the receipt of any antidepressant, as well as by category and substance. Lifetime antidepressant use was calculated as the proportion of individuals in receipt of a prescription at any point in time prior to the end of the calendar year, among those with full follow-up for that calendar year. The annual new prescription rates were calculated as the proportion of individuals in receipt of a prescription in a calendar year, among those with full follow-up for that calendar year and no antidepressant prescriptions in the previous calendar year. 95% confidence intervals for proportions were calculated using the Agresti-Coull method [21].

#### Indication recording around SSRI initiation

To assess indications for antidepressant use among adults starting an SSRI for the first time, we calculated the proportion with records of mental health conditions in the 60 days either side of the prescription. This analysis was restricted to SSRIs, the most common class, because the inclusion of other classes, with broader indications, would limit the ability to interpret differences in indication recording. Indications were anxiety and fear-related disorders, depressive disorders, behaviours that challenge, or other possible mental health indications (post-traumatic and stress-related disorders; somatoform and dissociative disorders; obsessive-compulsive and related disorders; eating disorders; and personality disorders.) 95% confidence intervals were calculated using the Agresti-Coull method [21].

#### Mean and median doses of citalopram, fluoxetine and sertraline

We performed dose analysis on the three most commonly prescribed SSRIs: sertraline hydrochloride, fluoxetine hydrochloride, and citalopram hydrobromide. To calculate the mean daily dose prescribed within a group we first calculated an individual-level mean daily dose for that year (without weighting by prescription duration), then calculated the mean across each of these individual-level averages. 95% confidence intervals for the mean doses were calculated using the t-distribution [22]. As doses were not normally distributed, we also calculated individual-level median doses in each year and used these to calculate median daily doses prescribed and interquartile ranges for each comparator group. As a sensitivity analysis to assess differences in maintenance doses prescribed, we repeated both mean and median calculations after keeping only individuals that also had the drug prescribed in the prior year.

#### Treatment duration

In duration analyses we calculated the proportion of antidepressant prescription courses that continued for 1 year, 2 years and 3 years or more. All antidepressant substances were considered together, so antidepressant switches were considered part of the same course. Both a ‘survival’ method and a proportion of patients covered (PPC) method were used [23]. In the survival method, prescription courses were derived for each patient by combining overlapping prescription records, including a 90 day permissible gap to allow for late prescription refills and missing duration information. Courses were assigned to each year by their start date. The Kaplan-Meier method was used to estimate the cumulative probability of treatment discontinuation over time for each comparator group, with censoring when patients left the cohort. 95% confidence intervals analysis were calculated with a log-transformation of the survival function. The PPC method is described in supplementary methods.

#### Stratified analyses

Each of the above analyses were repeated with the cohort stratified by sex. Age-stratified analysis was also performed with four age groups: 16-29 years old, 30-44 years old, 45-64 years old, and 65 years old and over (an age group that was not included in primary analyses).

Analyses were performed in R 4.5.1 and RStudio 2025.5.1. Code lists and scripts are available at https://github.com/awssadik/antidepressants-autism-uk.

## Results

### Cohort characteristics

34,345,537 people met the criteria to enter the study, including 172,242 autistic adults of whom 47,011 (27.3%) had a record of intellectual disability (ID) and 125,231 who did not (see study flowchart Figure S1). Autistic adults were more likely to be male, under 30 years old, have white ethnicity and have a history of mental health conditions (Table 1). The differences in ethnicity and conditions are possibly partly explained by the fact that non-autistic adults were more likely to enter the cohort in earlier calendar years, when there was less complete recording of ethnicity and health data, however these differences remained when the cohort was restricted to the start, middle and end of the study period: 1997, 2010 and 2023 (supplementary table 2).

From the 34,345,537 non-autistic adults that were included in the unmatched analyses, 188,029, 500,757 and 683,880 matched comparators were identified for the autism with ID, autism without ID and all autism groups, respectively. These comparators were well matched for year of birth, sex and cohort entry year, however average follow-up time was longer in autistic groups than non-autistic groups (supplementary table 3), leading to some differences in sex and age distribution at the calendar year level (supplementary table 4).

**Table 1:** Cohort characteristics, including all adults eligible to be included in the study, stratified by autism status. ADHD = attention deficit hyperactivity disorder; Psych contact = contact, referral or signposting to to a mental health service; IQR = interquartile range.

|  | Not autistic | Autistic without ID | Autistic with ID |
| --- | --- | --- | --- |
| People | 34173295 | 125231 | 47011 |
| Gender |  |  |  |
| Male | 16677089 (48.8%) | 85841 (68.5%) | 34958 (74.4%) |
| Female | 17495279 (51.2%) | 39281 (31.4%) | 12037 (25.6%) |
| Cohort entry year |  |  |  |
| 1997-2005 | 15333935 (44.9%) | 8406 (6.7%) | 5691 (12.1%) |
| 2006-2014 | 8585055 (25.1%) | 26224 (20.9%) | 13817 (29.4%) |
| 2015-2023 | 10066959 (29.5%) | 83322 (66.5%) | 26142 (55.6%) |
| Cohort entry age |  |  |  |
| 16-29 years | 13990078 (40.9%) | 107622 (85.9%) | 37302 (79.3%) |
| 30-44 years | 9757377 (28.6%) | 13035 (10.4%) | 6071 (12.9%) |
| 45-64 years | 6465995 (18.9%) | 4109 (3.3%) | 3189 (6.8%) |
| 65+ years | 3959845 (11.6%) | 465 (0.4%) | 449 (1%) |
| Ethnicity |  |  |  |
| White | 17826058 (52.2%) | 94164 (75.2%) | 34813 (74.1%) |
| South Asian | 2176954 (6.4%) | 2712 (2.2%) | 2603 (5.5%) |
| Black | 1169243 (3.4%) | 2728 (2.2%) | 2564 (5.5%) |
| Other | 990388 (2.9%) | 1263 (1%) | 612 (1.3%) |
| Mixed | 440107 (1.3%) | 2898 (2.3%) | 1083 (2.3%) |
| Unknown | 11570545 (33.9%) | 21466 (17.1%) | 5336 (11.4%) |
| Region |  |  |  |
| North West | 5803968 (17%) | 23535 (18.8%) | 8477 (18%) |
| North East | 1135931 (3.3%) | 5090 (4.1%) | 1925 (4.1%) |
| Yorkshire and The Humber | 1139642 (3.3%) | 3857 (3.1%) | 1339 (2.8%) |
| East Midlands | 940039 (2.8%) | 3502 (2.8%) | 887 (1.9%) |
| West Midlands | 5063357 (14.8%) | 21723 (17.3%) | 8835 (18.8%) |
| East of England | 1218551 (3.6%) | 4056 (3.2%) | 1593 (3.4%) |
| London | 7605825 (22.3%) | 15616 (12.5%) | 8128 (17.3%) |
| South East | 7614118 (22.3%) | 34862 (27.8%) | 11114 (23.6%) |
| South West | 3570306 (10.4%) | 12524 (10%) | 4582 (9.7%) |
| Northern Ireland | 81558 (0.2%) | 466 (0.4%) | 131 (0.3%) |
| Clinical |  |  |  |
| ADHD | 190916 (0.6%) | 25825 (20.6%) | 8944 (19%) |
| Depression | 7397190 (21.6%) | 49310 (39.4%) | 12617 (26.8%) |
| Anxiety | 5305901 (15.5%) | 51281 (40.9%) | 15578 (33.1%) |
| Self harm | 1634130 (4.8%) | 26743 (21.4%) | 10031 (21.3%) |
| Psych contact | 4773321 (14%) | 88433 (70.6%) | 33113 (70.4%) |
| Median follow-up in years (IQR) | 5 (1.8, 12.2) | 3.2 (1.2, 7) | 5 (2, 10.3) |

### Annual, lifetime, and new antidepressant prescriptions

Annual antidepressant prescribing increased from 5.1% to 14.7% for non-autistic adults between 1997 and 2023 and from 10.8% to 30.0% for autistic adults (Figure 1, supplementary table 5). Prescribing was consistently higher for autistic adults without ID than those with ID (Figure 2, supplementary table 5). In 2023, 28.0% of autistic adults with ID and 30.7% of autistic adults without ID were prescribed antidepressants, compared to 8.8% and 7.6% of their respective matched non-autistic comparators.

Lifetime antidepressant use also increased across between 1997 and 2023, from 9.5% to 33.2%% for non-autistic adults and 17.1% to 45.8%% for autistic adults (Figure 1, supplementary table 6). In 2023, 41.1% of autistic adults with ID and 47.6% of autistic adults without ID had ever been prescribed antidepressants, compared to 19.6% and 16.2% in their matched groups, respectively (Figure 2, supplementary table 6).

The proportion of adults starting an antidepressant having not been prescribed one in the previous calendar year peaked for all groups before 2019 and has decreased since (Figure 1, supplementary table 7). In 2023, this proportion was 3.9% for non-autistic adults and 7.1% for autistic adults (8.0% of those without ID and 5.0% of those with ID). Among the matched comparator groups, the 2023 values were 3.0% and 2.9% for autistic adults with ID, autistic adults without ID, respectively (Figure 2, supplementary table 7).

SSRIs were consistently the most frequently prescribed class of antidepressant for a ll groups (Figure S2, supplementary table 8), accounting for most of the increase in prescribing since 1997. In 2023, 22.9% of autistic adults with ID, 23.9% of autistic adults without ID and 10.1% of non-autistic adults were prescribed SSRIs. These values were 6.5% and 5.9% for matched comparators to autistic adults with ID and autistic adults without ID, respectively. Prescribing rates of SNRIs peaked in around 2003/4 for all groups before dropping and gradually increasing again towards the end of that decade. This pattern has been mostly driven by venlafaxine prescribing (supplementary table 9). Prescriptions for “other” antidepressants have increased consistently since 1997, which is largely attributable to mirtazapine (supplementary tables 8 & 9).

### Indications, doses and durations

Across all groups, prior to the year 2022 recording of depressive disorders has consistently been higher than recording of anxiety and fear-related disorders at the time of starting SSRIs (Figure 1, Figure 2, supplementary table 10). However, while recording of depressive disorders have plateaued or dropped since 2005, recording of anxiety and fear-related disorders have increasedl. In 2023, 51.6% of non-autistic adults had a depressive disorder recorded at the time of starting SSRIs and 44.0% had an anxiety or fear-related disorder (Figure 1). For autistic adults without ID, 41.2% had a depressive disorder and 49.7% an anxiety or fear-related disorder (Figure 2). For autistic adults with ID, 29.7% had a depressive disorder and 29.8% an anxiety or fear-related disorder. These values reflect a consistent pattern of anxiety and depression recording being lower for autistic adults with ID than the other two groups. Values for the matched comparator groups were all similar to the unmatched non-autistic group.

Average doses of fluoxetine, citalopram and sertraline generally increased across the study period and were higher for autistic adults than non-autistic across (Figure 1, Figure 2, supplementary tables 11 & 12). The median dose of sertraline for autistic adults was 50mg (IQR: 50-100mg) in 2000 and 100mg (IQR:50-120mg) in 2023, whereas the corresponding figures for non-autistic adults were 50mg (IQR: 50-50mg) in 2000 and 50mg (IQR: 50-100mg) in 2023 (Figure S5, supplementary table 12). These patterns were similar when the analyses were restricted to those who also had prescriptions in the prior calendar year (Figure S4, Figure S6, supplementary tables 13 & 14).

The proportion of adults continuing to take antidepressants 1, 2, and 3 years after initiation increased over time for both autistic adults and non-autistic adults (Figure 1, Figure 2, supplementary table 15). The values were consistently highest for autistic adults. For example, 52.5% of autistic adults with ID and 46.1% of autistic adults without ID starting antidepressants in 2020 continued them for at least 1 year. These figures were 33.6% and 33.5% in the respective matched comparator groups. These patterns were also seen when using the “proportion of patients covered” method (Figure S3, supplementary table 16).

**Figure 1.**
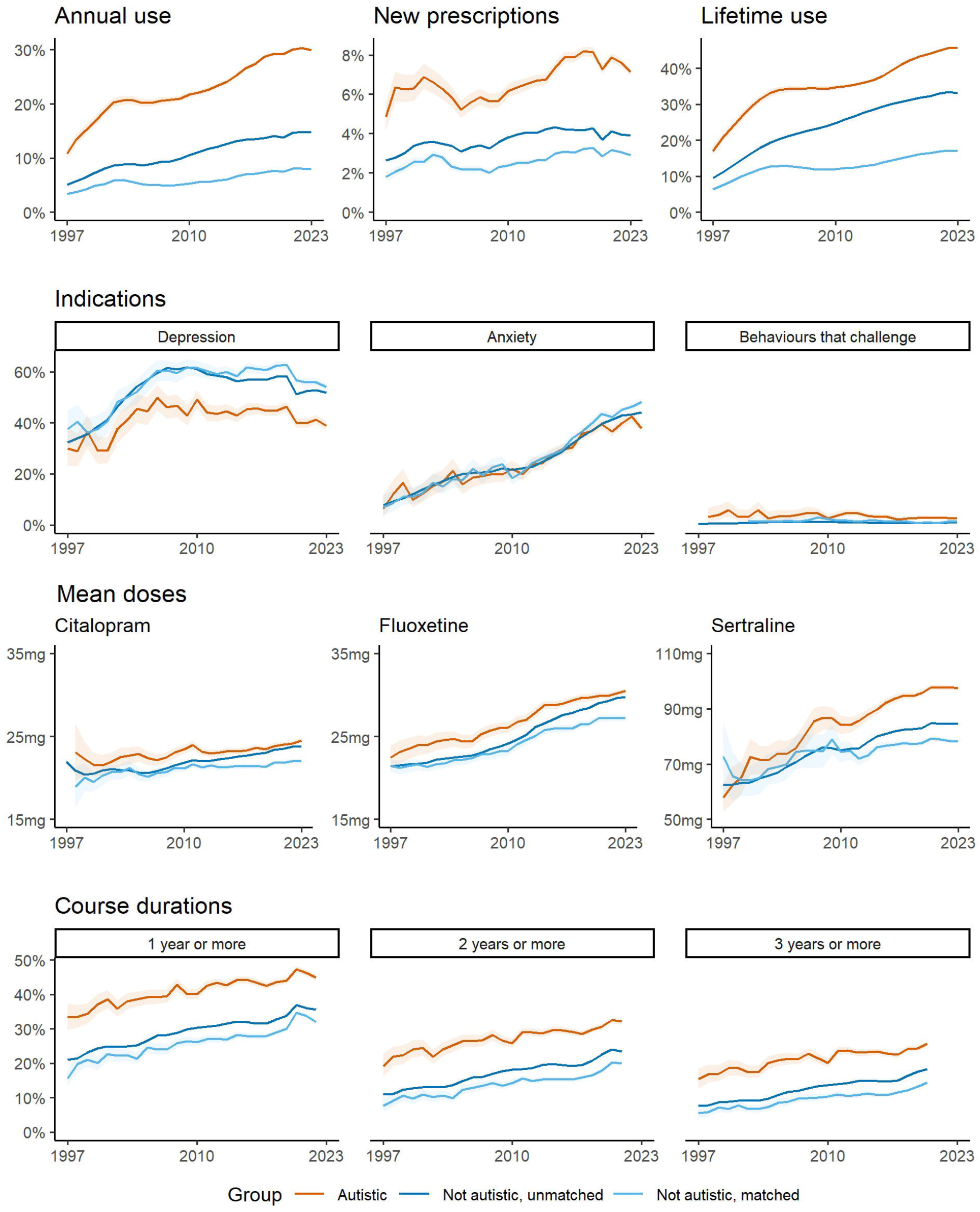
Antidepressant prescribing patterns for autistic adults, non-autistic adults, and matched non-autistic adults. Top row: proportion of adults prescribed antidepressants in each calendar year by autism status. (i) Any prescription in the current calendar year; (ii) New prescriptions among those without prescriptions in prior calendar year, (iii) Lifetime prescriptions prior to and including the current calendar year. Second row: proportion of adults starting an SSRI who have mental health conditions recorded in the two months either side, by calendar year and autism status. Third row: Mean dose of citalopram, fluoxetine and sertraline prescribed in each calendar year, by autism status. Bottom row: Proportion of antidepressant courses lasting at least 1 year, 2 years and 3 years, by year of course initation and autism status. Shaded areas represent 95% confidence intervals.

**Figure 2.**
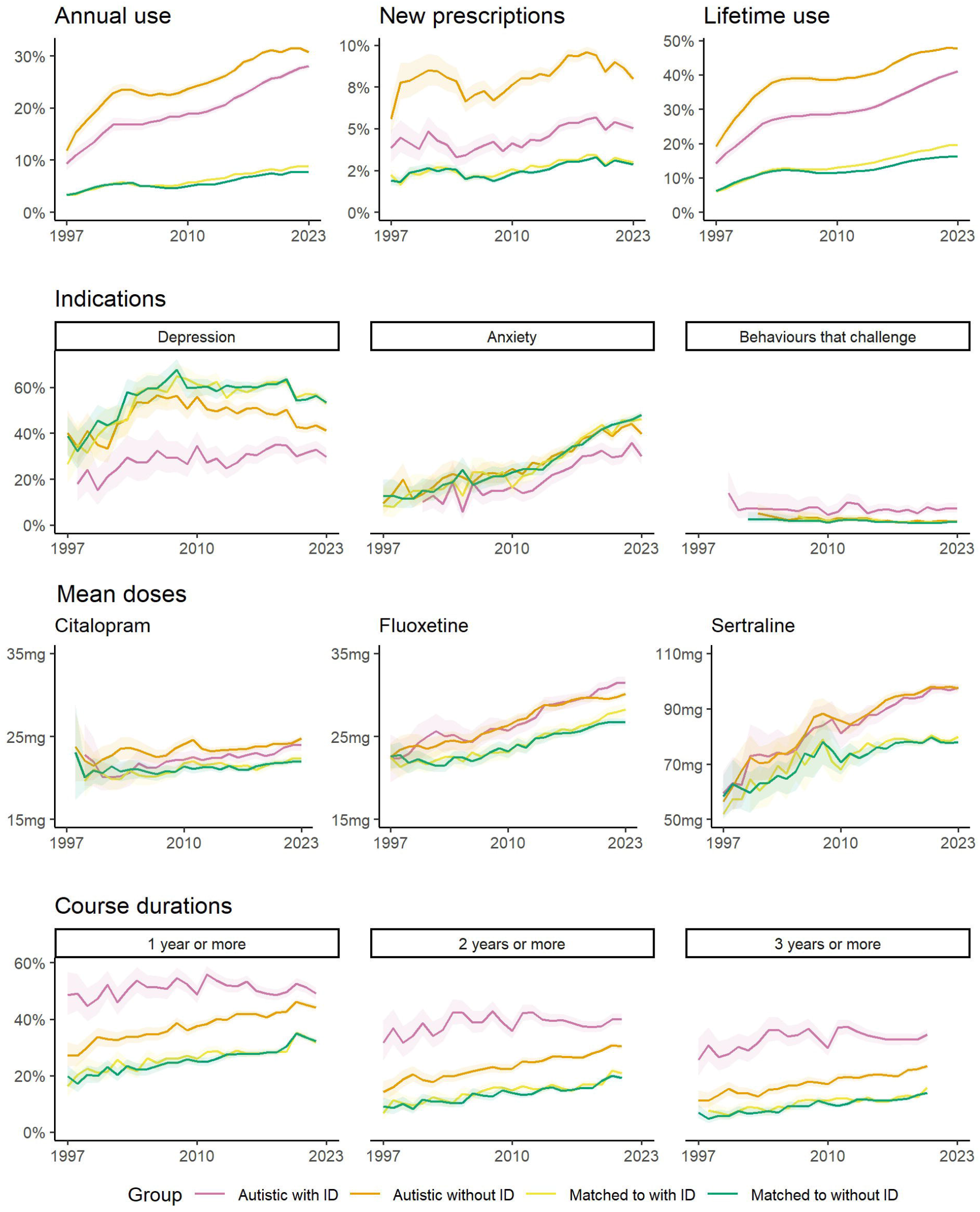
Antidepressant prescribing patterns for autistic adults with intellectual disability ID, autistic adults without ID, and matched non-autistic groups. Top row: proportion of adults prescribed antidepressants in each calendar year by autism status. (i) Any prescription in the current calendar year; (ii) New prescriptions among those without prescriptions in prior calendar year, (iii) Lifetime prescriptions prior to and including the current calendar year. Second row: proportion of adults starting an SSRI who have mental health conditions recorded in the two months either side, by calendar year and autism status. Third row: Mean dose of citalopram, fluoxetine and sertraline prescribed in each calendar year, by autism status. Bottom row: Proportion of antidepressant courses lasting at least 1 year, 2 years and 3 years, by year of course initation and autism status. Shaded areas represent 95% confidence intervals.

### Sex and age stratification

Sex stratified analyses showed higher overall prescribing and new prescribing rates among females compared to males, across all groups (Figure S7). Trends and patterns otherwise largely reflected those seen in the primary analyses.

Age stratified analyses also largely corresponded to the overall trends observed, although the uncertainty in results for over 65 year olds was high (Figure S8). Overall prescribing rates were lowest for 16-29 years olds across the comparator groups. New prescribing rates in the autistic without ID group were also lowest for 16-29 year olds, whereas in the autistic with ID group new prescribing rates were similar across age groups.

## Discussion

In this large cohort study using UK primary care data of over 34 million patients from 1997 to 2023, we found that antidepressant prescribing, doses, and duration of use have increased over time for both autistic and non-autistic adults. Furthermore, autistic adults had consistently higher proportions receiving new and ongoing antidepressant prescriptions, in higher doses and for longer durations than non-autistic adults. The proportions receiving ongoing and new prescriptions were highest for those without intellectual disability, whereas long term prescribing was more common for those with intellectual disability. Autistic adults with intellectual disability also had lower recording of anxiety and depression at antidepressant initiation compared to the other groups.

To our knowledge this is the largest study of antidepressant prescribing trends in autism and first to focus on adulthood. A previous study of psychotropic prescribing in UK primary care using The Health Improvement Network (THIN) database reported 3-4% of autistic people of all ages to be prescribed antidepressants between 1999 and 2008, with little change during this period [9]. A subsequent study using the same database reported ∼6% prescribed antidepressants in 2009 increasing steadily to 11-12% in 2016 [8]. By including children and adults in the same study, these studies may have understated the overall prescribing of antidepressants in the adult autistic population, where antidepressant prescribing is more common. We show similar trends of relatively steady levels of prescribing among autistic adults from 2001 to 2008, followed by a large increase between 2009 and 2016. We additionally show that this increase has continued since 2016, despite the launch of NHS England’s Stopping over-medication of people with a learning disability and autistic people (STOMP) initiative [6]. This may reflect the view that antipsychotics and dependency-forming medications are greater sources of potential harms, with less focus on antidepressants.

A recent study of the effect of the COVID-19 pandemic on antidepressant prescribing in the OpenSafely database of English primary care practices showed a brief drop in prescribing in March and April 2020 for the whole population (including for autistic people and those with ID), followed by a return to the trend of increasing prescribing rates [10]. Our findings also include a drop in prescribing rates in 2020 for autistic adults without ID and non-autistic adults. Additionally, our inclusion of data for 2023 suggests that prescriptions for autistic adults with ID and non-autistic adults have continued to increase, whereas this does not appear to be the case for autistic adults without ID.

Our observation of higher prescribing among females across autistic and non-autistic groups is also consistent with previous studies in the UK [8] and USA [24] [25] [26]. The finding that anxiety and depression recording rates among those starting antidepressants are similar between the two sexes suggests that the prescribing difference is primarily due to differences in co-occurring conditions [11]. While previous studies of antidepressant use in autistic adults are limited, our results for non-autistic adults are largely consistent with previous analyses in UK primary care datasets regarding indications [27] and length of prescriptions [5]. Our finding that doses prescribed of sertraline, fluoxetine and citalopram have increased over time for each group, is in keeping with previously reported trends towards increased prescribing of higher strength forms of sertraline and citalopram from 1998 to 2018 [28]. However, a study of prescribing in NHS Greater Glasgow and Clyde reported higher mean doses than at the same period of our study [29]. This may reflect differences in study methods or clinical practice.

Strengths of this study are its large size and the low risks of recall bias and selection bias, due to the use of prospectively routinely recorded information and inclusion of the all eligible patients from population-representative data.

Study limitations include the possibility of incomplete recording of conditions, doses and durations in primary care records which may lead to measurement bias. When stratifying the study population by autism and intellectual disability, we mitigated the impact of possible changes over time in record completeness by defining groups by any lifetime diagnoses, rather than requiring a diagnosis prior to entry to the group.

Regarding indications, our finding of an increased proportion of antidepressant starters with anxiety is consistent with prior reports from UK primary care records and clinicians [30]. For duration analyses, the possible impact of missing information was mitigated by using a large 90 day permissible gap in primary analyses, so all prescriptions were considered at least 90 days long. Another study limitation is that the matched comparators were not necessarily balanced within each calendar year, because of differential follow-up between groups. This is unlikely to affect the study conclusions, given that differences between groups were consistent in the matched and unmatched analyses. Finally, secondary care prescribing information was unavailable, however antidepressant prescriptions for adults in the UK are generally made in primary care even when recommended by a secondary care clinician.

Our findings of higher antidepressant prescribing for autistic adults may reflect increased mental health conditions and/or off-label use. For those without ID, the proportion of SSRI initiators with recorded depression or anxiety are similar to the non-autistic adults, suggesting that higher prescribing mostly reflects appropriate prescribing in the context of higher anxiety and/or depression rates. This is not the case for those with ID, suggesting off-label prescribing has a greater role.

The higher doses of SSRIs prescribed to autistic adults could be due a greater proportion of autistic adults being on maintenance doses, however, the differences remained when analyses were restricted to those who also had prescriptions in the prior calendar year. Alternatively, the higher doses may reflect more severe anxiety or depression and a lack of therapeutic response to lower doses, or a reluctance of prescribers or patients to reduce dose after a period of stability. This latter explanation may also explain greater long-term use among autistic adults.

## Conclusions

This study confirms that antidepressant prescribing has continued to increase in the UK and that they were prescribed and initiated more frequently among autistic adults, particularly females and those without intellectual disability. Similarly, autistic adults continued to be prescribed antidepressants for longer and were prescribed higher doses of SSRIs on average than non-autistic adults.

To build on these findings, cross-nation comparisons and studies of other psychotropic medications are required to estimate the generalisability of results and full impact of deprescribing initiatives; mixed methods studies are required to unpick the reasons for higher antidepressant initiation and lower discontinuation rates; and randomised controlled trials are needed to establish the therapeutic effectiveness and side effect profiles in autistic adults [31].

## Supporting information

RECORD-PE Checklist

## Data Availability

This study is based in part on data from the Clinical Practice Research Datalink obtained under licence from the UK Medicines and Healthcare products Regulatory Agency. The data is provided by patients and collected by the NHS as part of their care and support. The interpretation and conclusions contained in this study are those of the author/s alone. Instructions for how to access the data are available at https://www.cprd.com/access-data.
Analytic code are available at https://github.com/awssadik/antidepressants-autism-uk.

## Acknowledgements

AS would like to thank his PhD Advisory Group members L Ting and Ben Argo for their feedback throughout the research cycle.

## Author Contribution

AS: formulating questions, performing analyses, study design, writing article

PMD, DR: formulating questions, guidance on performing analyses, study design, writing article

GMK, AFP, BL, ML, CM: formulating questions, study design, writing article

## Transparency Declaration

The manuscript is an honest, accurate, and transparent account of the study being reported. No important aspects of the study have been omitted. Any discrepancies from the study as planned and registered have been explained.

## Data Availability

This study is based in part on data from the Clinical Practice Research Datalink obtained under licence from the UK Medicines and Healthcare products Regulatory Agency. The data is provided by patients and collected by the NHS as part of their care and support. The interpretation and conclusions contained in this study are those of the author/s alone. Instructions for how to access the data are available at https://www.cprd.com/access-data.

## Analytic Code Availability

Analytic code are available at https://github.com/awssadik/antidepressants-autism-uk.

## Research Material Availability

Variable code lists are available at https://github.com/awssadik/antidepressants-autism-uk.

