## Supplementary Figures for "Antidepressant prescribing trends for adults with and without autism in the United Kingdom from 1997 to 2023, a population-based cohort study using Clinical Practice Research Datalink Aurum"

Figure S1 : Participant flowchart

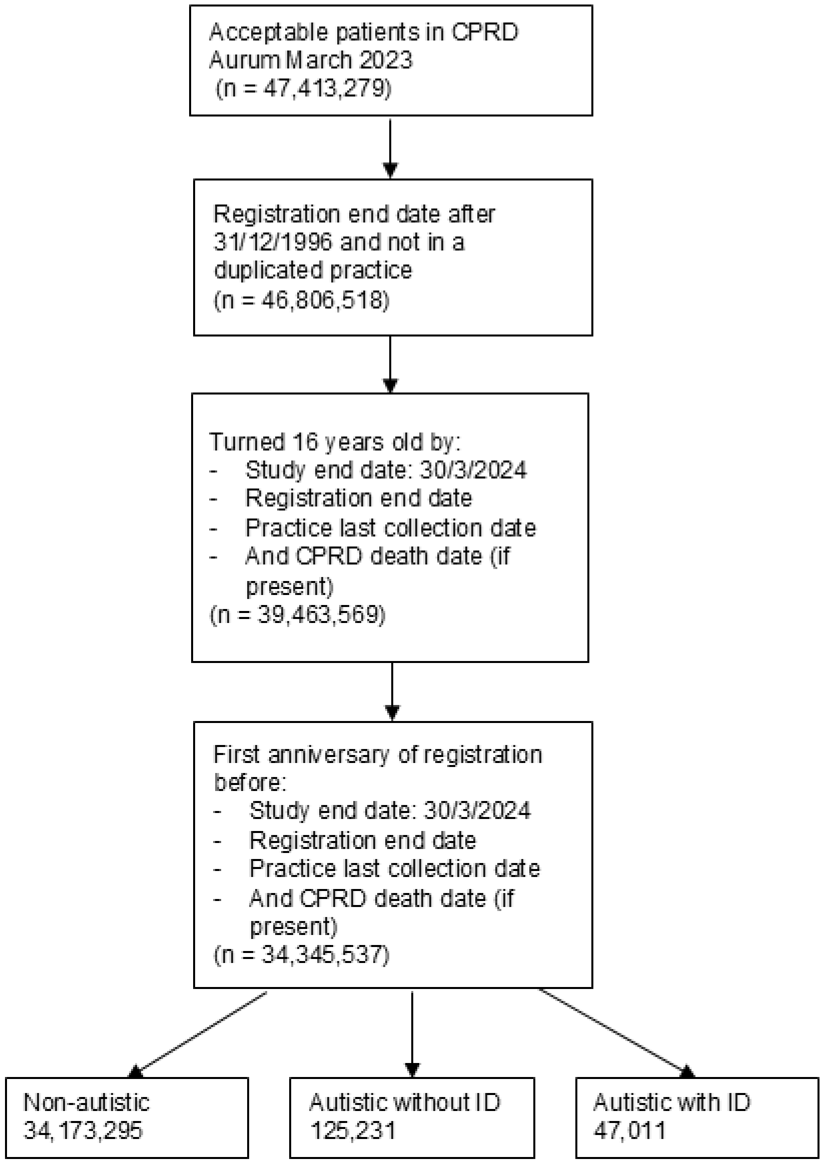


*Figure S1*

Figure S2 : Annual proportion of people prescribed each class of antidepressant by autism status. SSRI = serotonin-selective reuptake inhibitor; SSRI = serotonin and noradrenaline reuptake inhibitor; TCA = tricyclic antidepressant; MAO-I = monoamine-oxidase inhibitor. Shaded areas represent 95% confidence intervals.

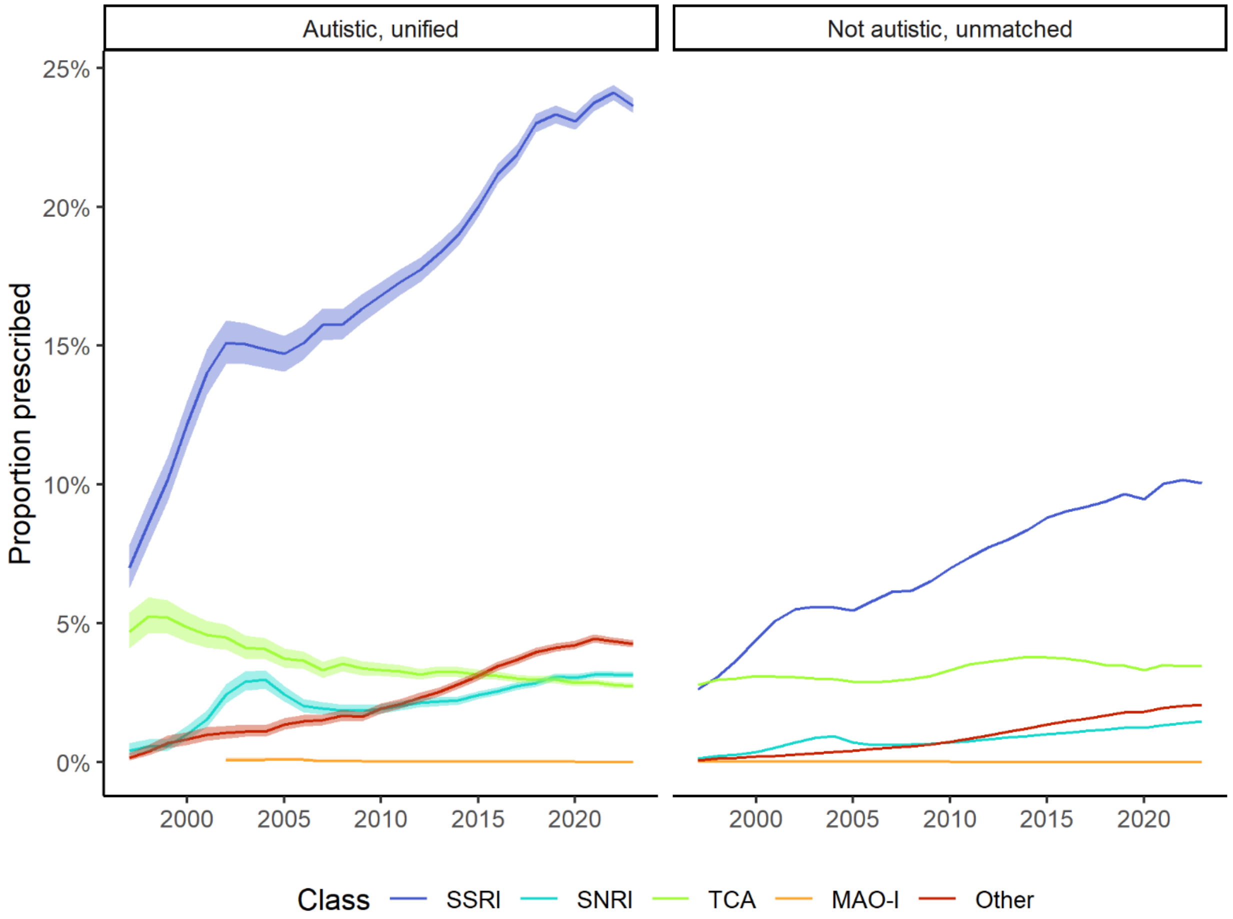


*Figure S2*

Figure S3 : Proportion of patients taking antidepressants at 1 year, 2 years and 3 years after starting an antidepressant, by calendar year and autism status. Shaded areas represent 95% confidence intervals.

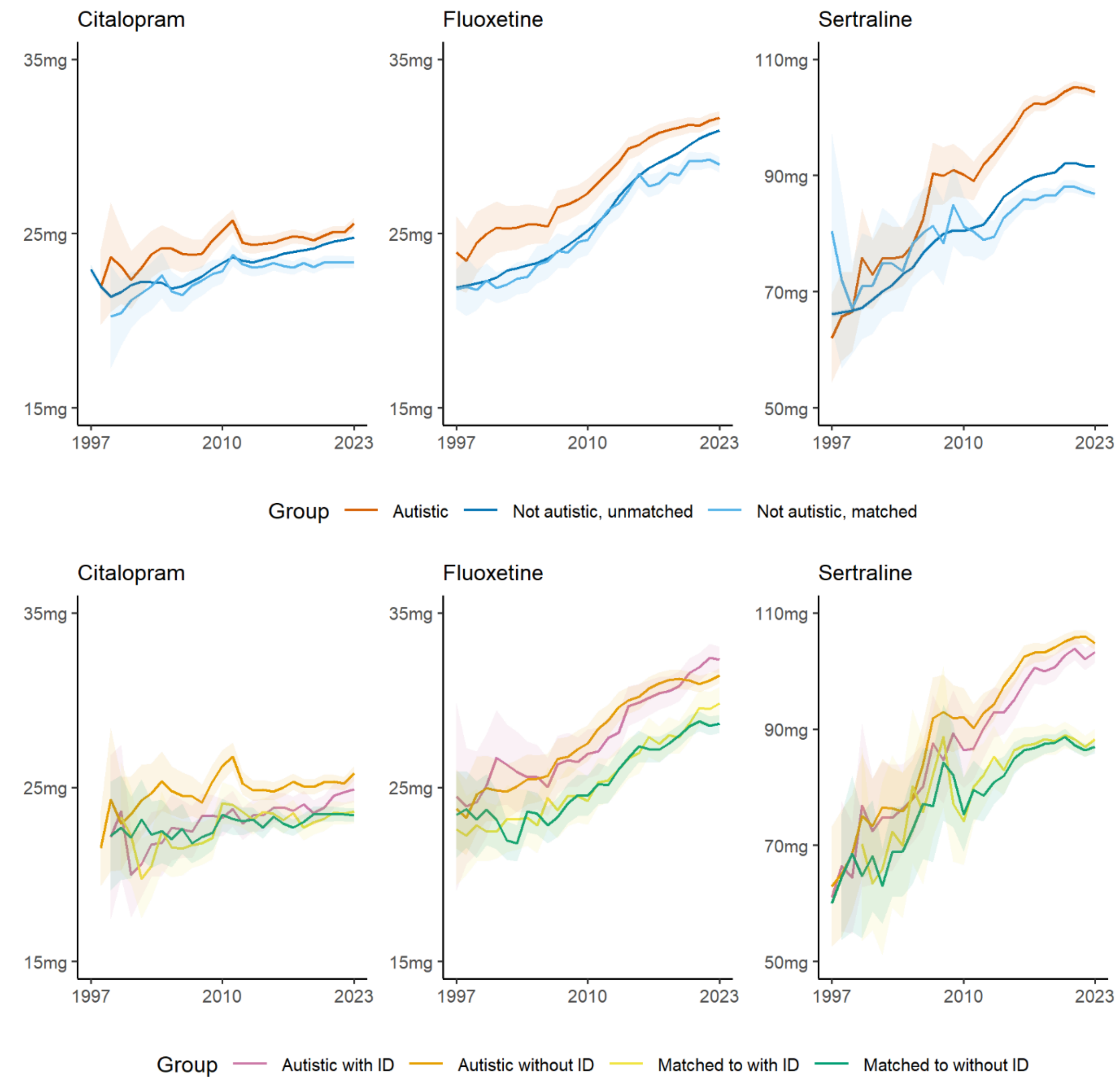


*Figure S3*

Figure S4 : Mean doses of citalopram, fluoxetine and sertraline prescribed in each calendar year, by autism status, after excluding patient-years in which antidepressants were being initiated. Shaded areas represent 95% confidence intervals.

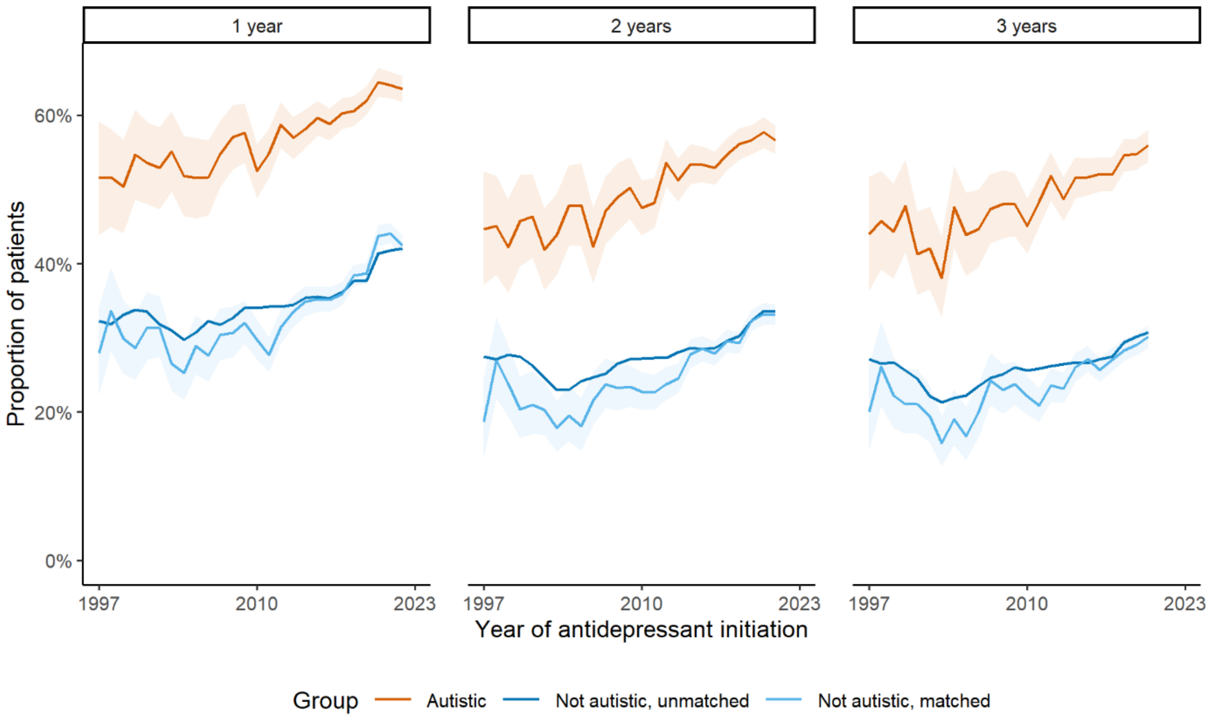


*Figure S4*

Figure S5 : Median doses of citalopram, fluoxetine and sertraline prescribed in each calendar year, by autism status. Shaded areas represent interquartile range.

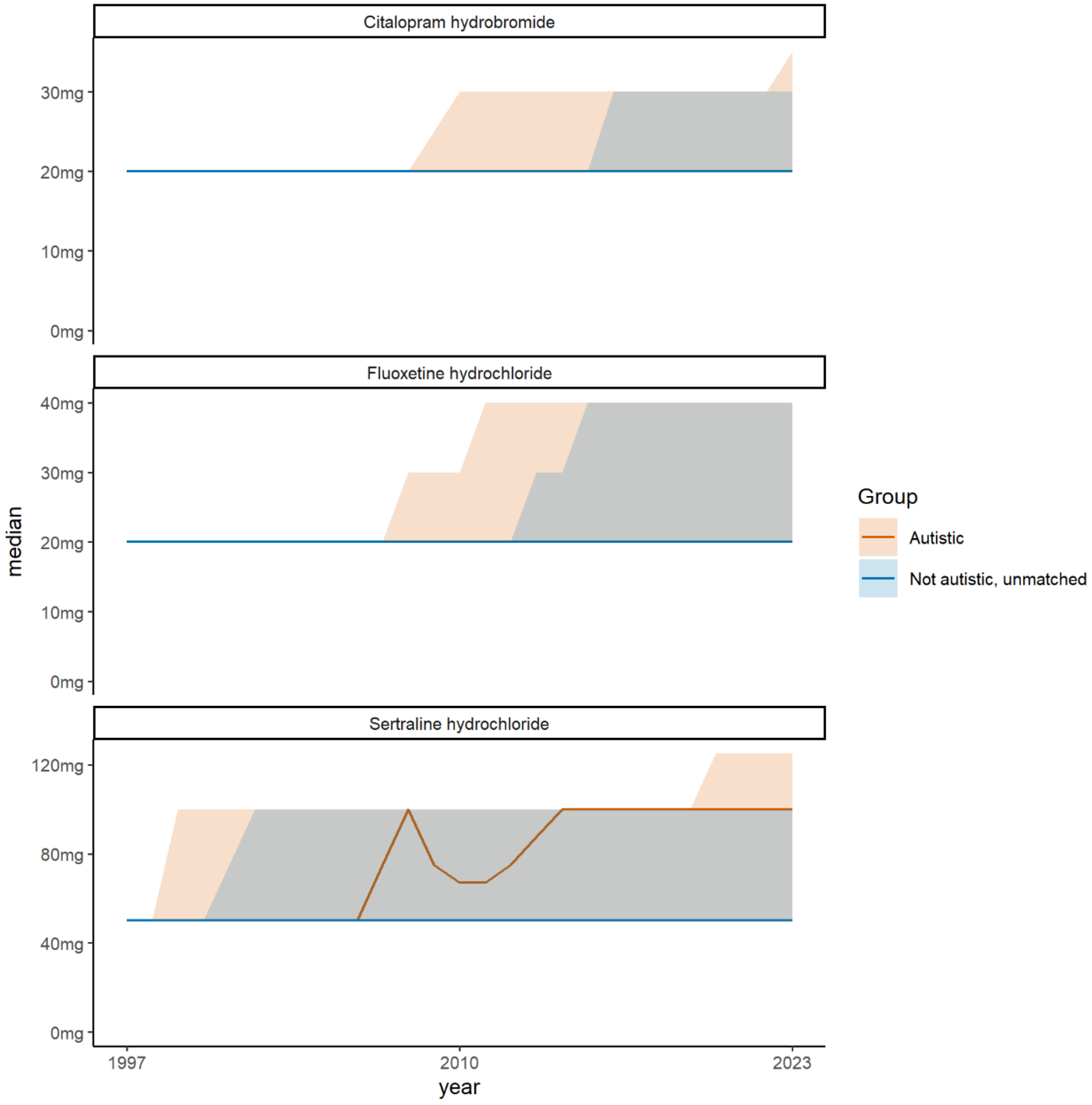


*Figure S5*

Figure S6 : Median doses of citalopram, fluoxetine and sertraline prescribed in each calendar year, by autism status, after excluding patient-years in which antidepressants were being initiated. Shaded areas represent interquartile range.

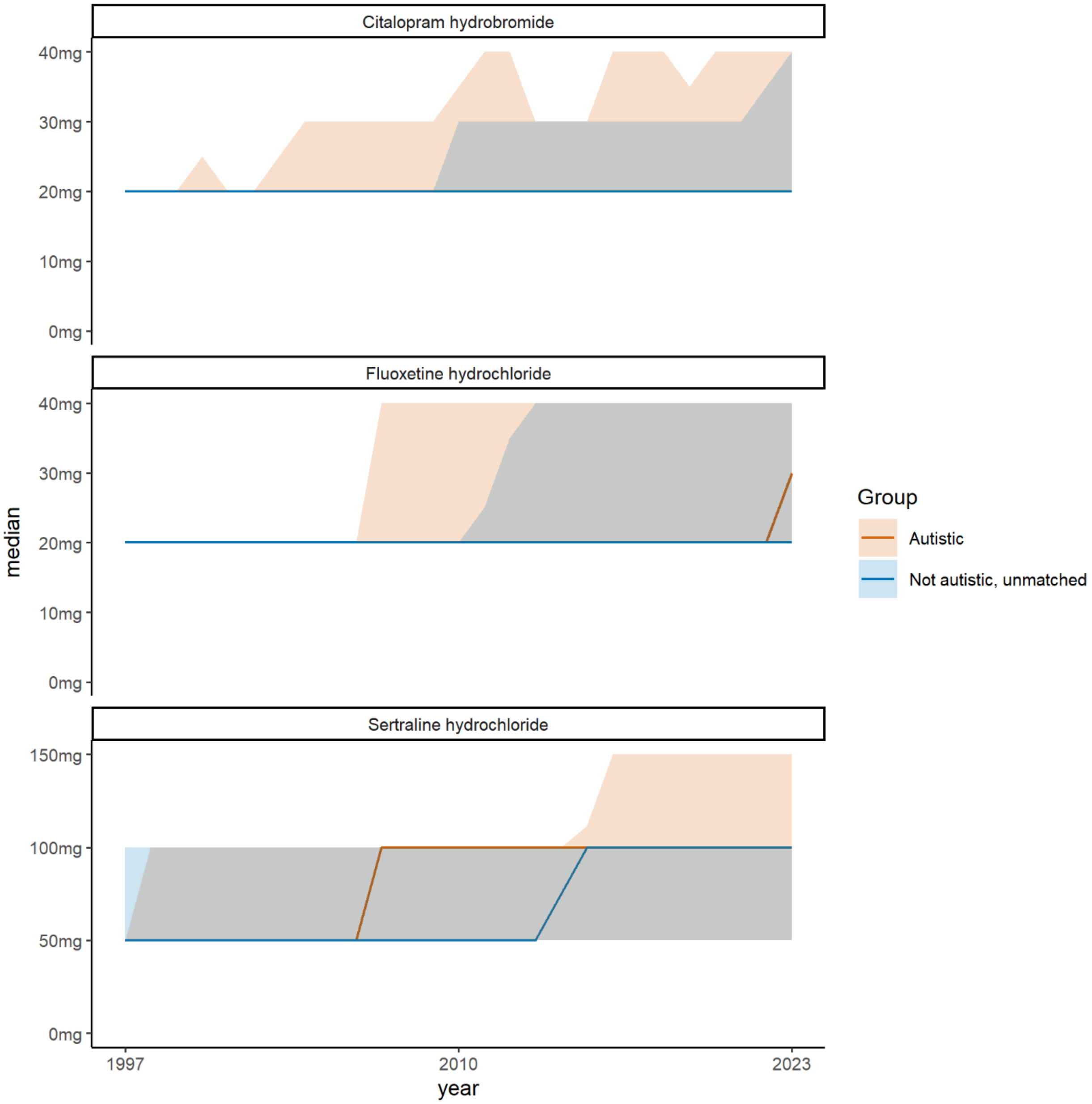


*Figure S6*

Figure S7: Antidepressant prescribing patterns for autistic adults with intellectual disability (ID), autistic adults without ID and unmatched non-autistic adults, by sex. Top row: proportion of adults prescribed antidepressants in each calendar year by autism status - any prescription in the current calendar year. Second row: Top row: proportion of adults prescribed antidepressants in each calendar year by autism status - new prescriptions among those without prescriptions in prior calendar year. Third & forth rows: proportion of adults starting an SSRI who have anxiety or depression recorded in the two months either side, by calendar year and autism status. Fifth row: Mean dose of sertraline prescribed in each calendar year, by autism status. Bottom row: Proportion of antidepressant courses lasting at least 2 years, by year of course initation and autism status. Shaded areas represent 95% confidence intervals.

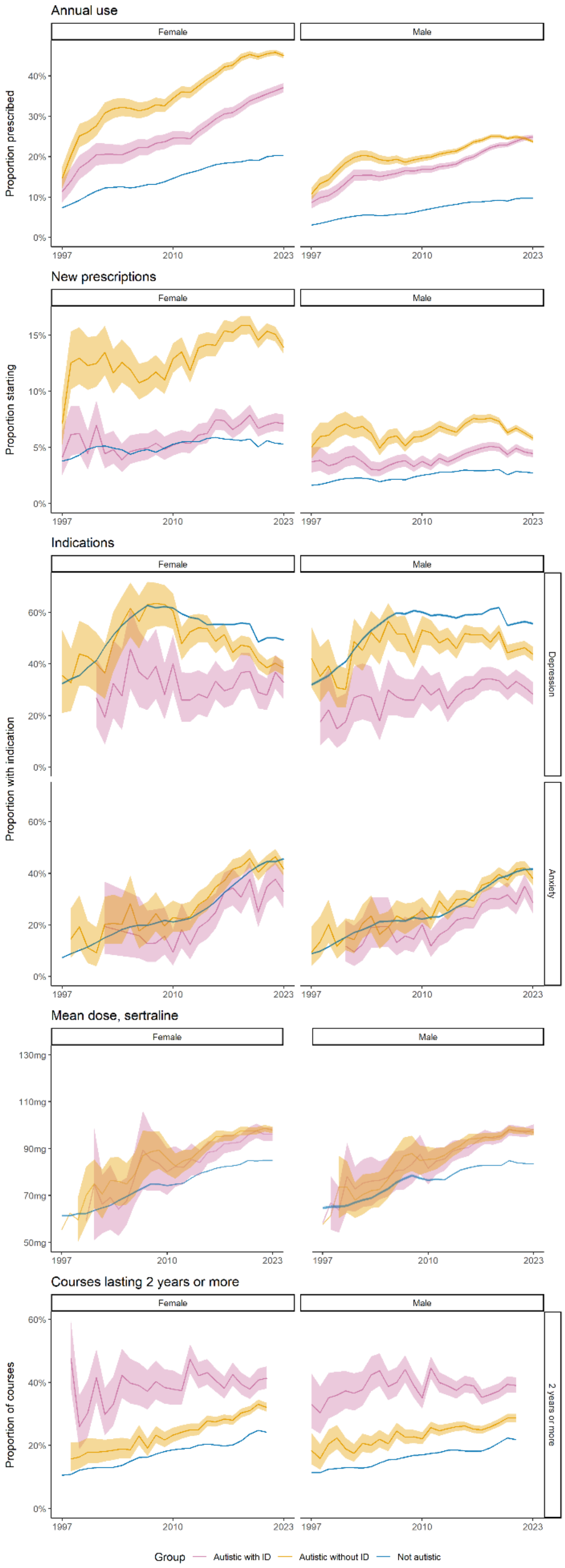
Annual use
Female
Male
Proportion prescribed
New prescriptions
Proportion starting
Indications
Proportion with indication
Depression
Anxiety
Mean dose, sertraline
Courses lasting 2 years or more
Proportion of courses
2 years or more
Group
Autistic with ID
Autistic without ID
Not autistic

Figure S8: Antidepressant prescribing patterns for autistic adults with intellectual disability (ID), autistic adults without ID and unmatched non-autistic adults, by age-group. Top row: proportion of adults prescribed antidepressants in each calendar year by autism status - any prescription in the current calendar year. Second row: Top row: proportion of adults prescribed antidepressants in each calendar year by autism status - new prescriptions among those without prescriptions in prior calendar year. Third & forth rows: proportion of adults starting an SSRI who have anxiety or depression recorded in the two months either side, by calendar year and autism status. Fifth row: Mean dose of sertraline prescribed in each calendar year, by autism status. Bottom row: Proportion of antidepressant courses lasting at least 2 years, by year of course initation and autism status. Shaded areas represent 95% confidence intervals.

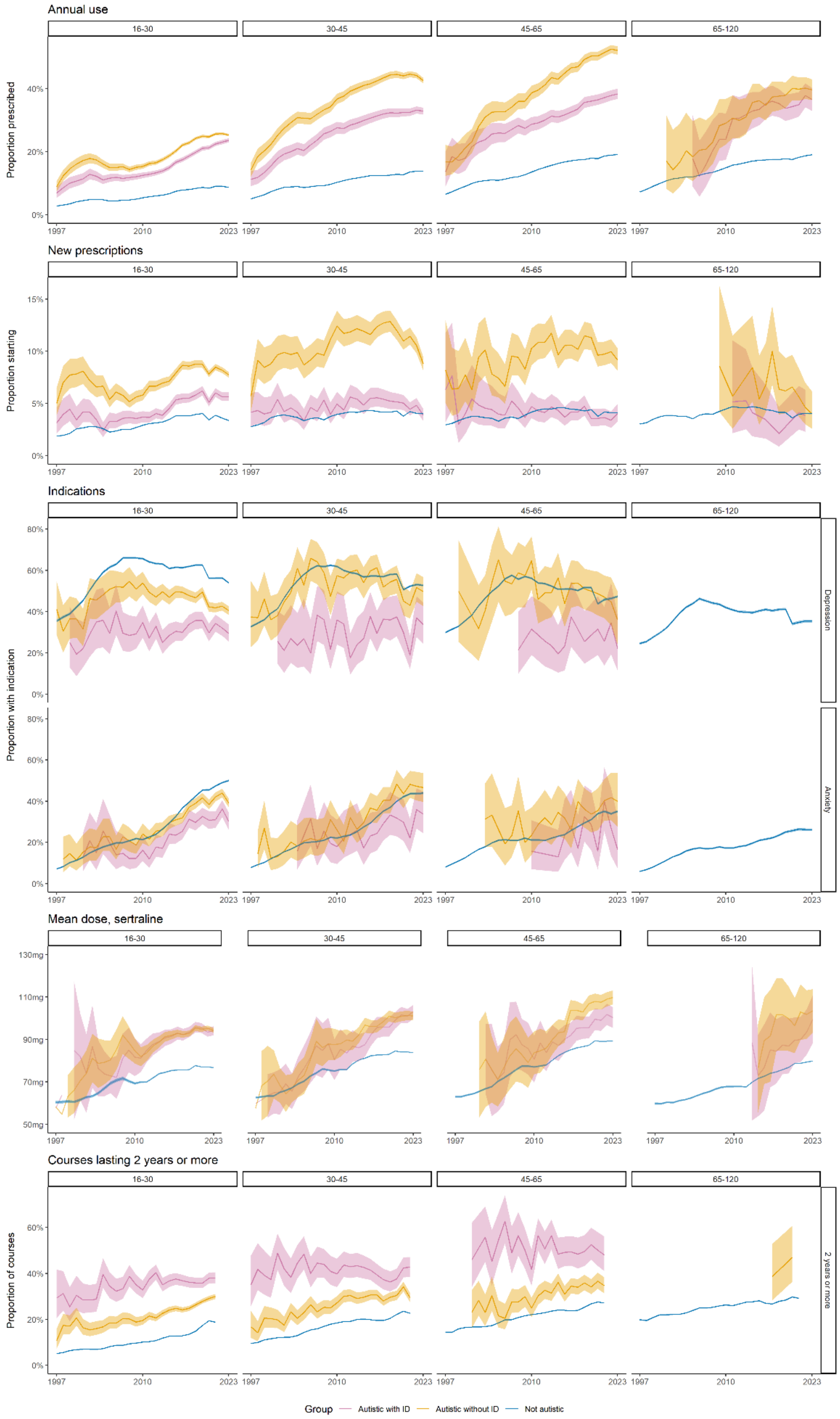

Annual use
16-30
30-45
45-65
65-120
Proportion prescribed
0%
20%
40%
1997
2010
2023
New prescriptions
Proportion starting
0%
5%
10%
15%
Indications
Proportion with indication
0%
20%
40%
60%
80%
Depression
Anxiety
Mean dose, sertraline
50mg
70mg
90mg
110mg
130mg
Courses lasting 2 years or more
Proportion of courses
2 years or more
Group
Autistic with ID
Autistic without ID
Not autistic
