## Supplementary Methods for "Antidepressant prescribing trends for adults with and without autism in the United Kingdom from 1997 to 2023, a population-based cohort study using Clinical Practice Research Datalink Aurum"

**Code lists development**

The CPRD Code Browser was used to prepare code lists. Search terms were determined after reviewing published code lists and current and historical psychiatric categorisation systems. Relevant codes were selected by their description. The SNOMED Concept IDs for these codes were then used to identify further codes for review. The code lists were then crosschecked against relevant lists from Health Data Research UK Phenotype Library[16], OpenCodelists [17], and published multimorbidity studies [18]. Codes present in at least one source were included in the final code list and all other codes were reviewed for inclusion. AS (a psychiatric resident doctor) carried out all the searches and reviewed search results against published code-lists. Where relevant, advice from another clinician (DR) was sought. For the cohort stratification variables - autism and intellectual disability - records without observation dates were not included, as these were considered to be potentially erroneous.

Ethnicity codes were developed by identifying published lists [[19]][20] and searching for additional codes in the CPRD Code Browser.

Antidepressant substances were classified with guidance from the British National Formulary (BNF) and NHS Business Services Authority's BNF codes list (version 2nd May 2024) [32]. The codes for each substance were identified by a wildcard search of the substance name in the CPRD Code Browser. Categories were serotonin-selective reuptake inhibitors (SSRIs), serotonin-noradrenaline reuptake inhibitors (SNRIs), tricyclic antidepressants (TCAs), monoamine oxidase inhibitors (MAO-I) and other antidepressants. Included antidepressants are listed in supplementary table 1.

The term *sex* is used throughout this study because the variable labelled *gender* in CPRD data generally reflects the sex recorded at birth, based on biological characteristics.

**Prescription dose cleaning**

If there were multiple drug issues of the same substance on the same date for the same person, these were combined into a single prescription and the duration and daily dose calculated as follows. First, the total strength of medication prescribed in milligrams was calculated. If this was not possible, the dose was set to missing. Then, to determine whether combined prescriptions should be considered to be concurrent or consecutive, both the maximum duration and the sum of the durations in the group were calculated. Concurrent prescriptions were deemed plausible if dividing total strength by the maximum duration led to a plausible daily dose for that substance. Consecutive prescriptions were deemed plausible if dividing total strength by the sum of durations led to a plausible daily dose for that substance. If both were plausible, then the duration information was used to determine the most plausible option based on the following ranking:

i) Concurrent if the combined daily dose for taking prescriptions at the same time is plausible, and the highest duration is 28 days or more
ii) Consecutive if the average daily doses are plausible for taking consecutively, and combined duration is 28 or less
iii) Otherwise prioritise concurrent before consecutive.

If neither concurrent nor consecutive prescriptions were deemed plausible, the duration was set to the maximum duration among the multiple drug issues and the daily dose was set as missing.

When there was a single prescription for a particular substance on a particular date for someone but the daily dose was missing or implausible, it was estimated based on other information: (i) multiplying single dose strength by daily dose frequency; (iii) dividing the total quantity prescribed by the cleaned duration of the prescription; or (iii) simply using the tablet strength as the daily dose (thus assuming one tablet would be taken daily). If none of this information was available the daily dose was set to missing.

**Treatment duration: proportion of patients covered method**

In the PPC method, patients who were starting an antidepressant for the first time were identified within each calendar year. For each patient, the index date was set as the date of this first prescription and we identified whether they were taking an antidepressant at 1 year, 2 years, and 3 years after this index date. The proportion of patients covered was calculated for each time point as the number of patients taking an antidepressant divided by the number of patients that remained in the cohort. So, in contrast to the survival method, patients who stopped taking an antidepressant could later still contribute to the numerator with subsequent prescription courses. The PPC approach is understood to be less sensitive to the gap length specified [23]. 95% confidence intervals were calculated using the Agresti-Coull method [21]

*Please see main article for references*.
